# A Rapidly Deployable Negative Pressure Enclosure for Aerosol-Generating Medical Procedures

**DOI:** 10.1101/2020.04.14.20063958

**Authors:** Anthony M. Chahal, Kenneth Van Dewark, Robert Gooch, Erin Fukushima, Zachary M. Hudson

## Abstract

**Background:** The coronavirus disease 2019 (COVID-19) pandemic presents significant safety challenges to healthcare professionals. In some jurisdictions, over 10% of confirmed cases of COVID-19 have been found among healthcare workers. Aerosol-generating medical procedures (AGMPs) may increase the risk of nosocomial transmission, exacerbated by present global shortages of personal protective equipment (PPE). Improved methods for mitigating risk during AGMPs are therefore urgently needed.

**Methods:** The Aerosol Containment Enclosure (ACE) was constructed from acrylic with silicone gaskets for arm port seals and completed with a thin plastic sheet. Hospital wall suction generated negative pressure within the ACE. To evaluate protective capability, differential pressures were recorded under static conditions and during simulated AGMPs. Smoke flow patterns, fluorescence aerosolization, and sodium saccharin aerosolization tests were also conducted.

**Results:** Negative pressures of up to -47.7 mmH_2_O were obtained using the enclosure with two wall suction units (combined outflow of 70 L min^-1^), with inflow of O_2_ of 15 L min^-1^. Negative pressures between -10 and - 35 mmH_2_O were maintained during simulated AGMPs, including oxygen delivery by mask, airway suctioning, bag-mask manual ventilation and endotracheal intubation of a potential COVID-19 patient. The ACE effectively contained smoke, fluorescein aerosol, and sodium saccharin aerosol within the enclosure during use.

**Conclusions:** The ACE is capable of maintaining negative pressure during simulated AGMPs. In all cases, containment was improved relative to an identical enclosure with non-occluded ports at ambient pressure. During the current COVID-19 pandemic, the use of such a device may assist in reducing nosocomial infections among healthcare providers.

## Introduction

Coronavirus disease 2019 (COVID-19) has created an unprecedented risk to both health care providers and populations, with a coordinated global response confounded by limited resources. At the forefront of this response, healthcare workers face challenges in obtaining adequate personal protective equipment (PPE), creating a need for innovative solutions to better protect them from infection.

Many patients with COVID-19 present with respiratory distress and require immediate airway support. Intubation, bag-mask ventilation (BMV), and the provision of high-flow oxygen may cause aerosolization of viral particles, increasing the risk to healthcare providers.^1^ During the Severe Acute Respiratory Syndrome (SARS) outbreak in 2003, intubation was identified as an independent risk factor for physician illness.^1^

Negative pressure rooms, minimizing bedside staff, and wearing appropriate PPE are all recommended to mitigate the risk to healthcare providers during AGMPs. Recommendations for intubation suggest limiting oxygen flows for preoxygenation, avoiding apneic oxygenation, and avoiding bag-mask ventilation. Modified procedures during intubation include the use of a plastic sheet to cover the patient, or the use of a solid plastic shield separating healthcare providers from the patient’s airway.^2^

The authors propose that a negative pressure enclosure over a patient during intubation could reduce the risk of virus transmission to the healthcare team during AGMPs. Building upon the open plastic shield first proposed by Dr. Hsien-Yung Lai, the authors developed a negative pressure micro-enclosure which limits outward aerosol transmission.^3^ This enclosure can be assembled at limited cost and deployed rapidly during emergent airway management, with simple resources found in many emergency departments and critical care spaces. Post-procedure, the enclosure can be cleaned for immediate reuse.

## Background

Healthcare providers are inherently at risk for nosocomial infections due to their close interactions with patients. During the SARS epidemic, 20% of all cases involved healthcare workers.^4^ Similarly, in

Zhongnan Hospital in Wuhan, China, between January 1st and 28th, 2020, 40 patients of the 138 admitted to the hospital with COVID-19 were healthcare workers.^5^ In Wuhan, by February 12, 2020, 3019 healthcare workers had been infected by SARS-CoV-2, accounting for approximately 4% of all cases.^6^ In Italy 12,252 healthcare workers have contracted COVID-19 as of April 5, 2020, representing 10.2% of all Italian cases.^7^ Similarly, as of April 2, 2020, in Ontario, Canada, 9.6% of all cases of COVID-19 have been reported in healthcare workers.^8^ The high rates of infection in healthcare staff likely plays a key role in stressing healthcare delivery through human resource depletion.^9^ Any means of minimizing these risks to healthcare delivery capacity is thus urgently required.

Transmission through infectious droplets and aerosols represent significant components of this risk, with unique PPE requirements needed to mitigate each. Droplets, respiratory particles > 5 μm in diameter, tend to travel distances of 1 meter or less and settle quickly, whereas aerosols, particulates < 5 μm in diameter, remain in the air for longer and travel further.^10^ This is of particular concern during the current COVID-19 pandemic as many AGMPs are potentially life-sustaining interventions for such infected patients. Unfortunately, aerosols containing SARS-CoV-2 may remain viable for several hours after they are produced.^11^

Negative pressure rooms are an important tool for mitigating infection risk from aerosolized particles.^10^ These rely on airflow from outside the environment into the closed room, which is then extracted through isolated vents, thereby safely containing aerosolized particles. This is akin to a biosafety cabinet, which uses airflow to protect personnel from their contents. National Sanitation Foundation / American National Standards Institute guidelines specify that Class II biosafety cabinets require a minimum flow of 0.38 - 0.51 m/s (75-100 ft/min) to contain aerosols.^12^ Air velocity through a gap is directly proportional to the airflow and inversely proportional to the area of the gap, as described below:

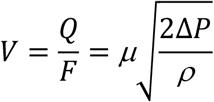

where *V* is air velocity (m/s), *Q* is airflow (m^3^/s), *F* is the gap area (m^2^), *µ* is the flow coefficient (1), Δ *P* is the pressure differential (Pa) and *ρ* is the air density (kg/m^3^).^13^

## Methods

The authors conceived the Aerosol Containment Enclosure (ACE) by combining a modified intubating shield and plastic sheeting to fully enclose a patient. Negative pressure was generated using two standard hospital suctions, each providing up to 260 mmHg of suction with 35 L min^-1^ flow. The ACE was constructed from a four-sided acrylic frame with silicone gaskets on the arm ports, at a total cost of approximately $200 USD. Two arm ports were created for the primary airway operator, and two for an airway assistant standing to the primary operator’s right.

Relative to the initial design reported by Dr. Lai, the arm ports were enlarged to 15.2 cm (6 in) to improve ergonomics, with 1.58 mm (1/16 inch) 30A durometer silicone gaskets added to provide an improved seal around the operators’ arms (Figure 1). All connections to the patient including monitor cables, intravenous lines, and oxygen tubing may be either run under the plastic sheet or under the enclosure. This allows for easy removal of the box without having to disconnect or withdraw any lines through the ports. Flow into the ACE occurs through small leaks around the patient and the perimeter of the enclosure, through the mattress cover, and through the patient’s oxygen source.

**Figure 1.**
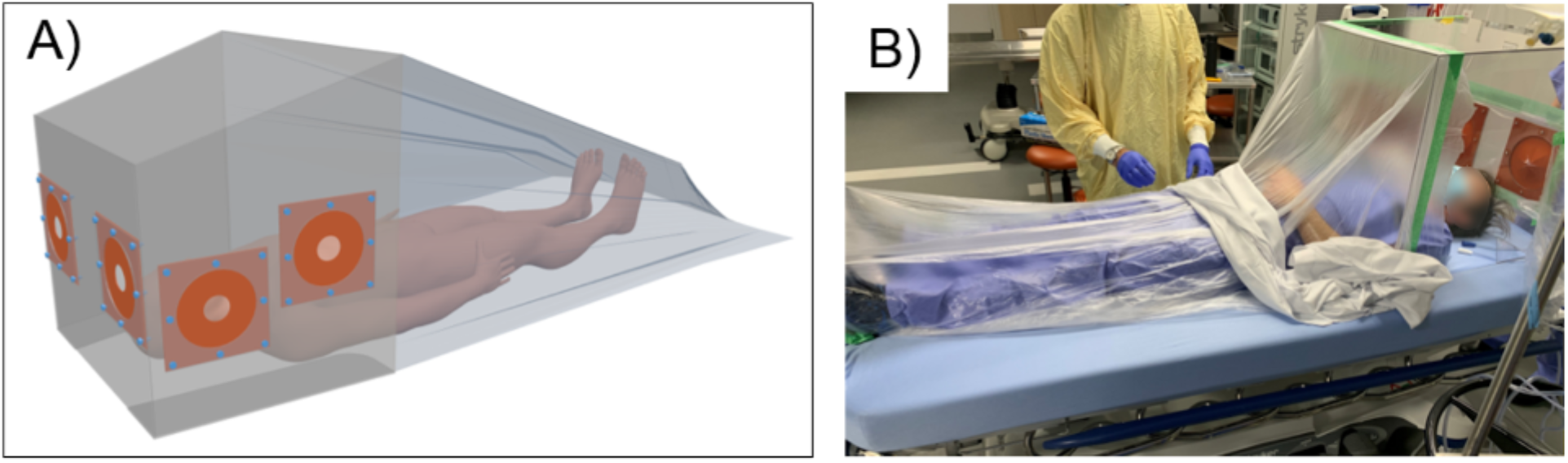
A) Schematic representation of the ACE in use with a plastic drape. B) Simulated clinical set-up of the ACE with a patient and negative pressure applied.

The caudal end of the box was enclosed using 0.008 mm (0.31 mil) high-density polyethylene (HPDE) plastic sheeting, chosen for its flexibility and strength. This material, designed for taping, draping and drop sheet covers, is readily available from painting supply stores. The extended width of this plastic is 2.7 m (9 ft), and it can be cut to length as desired, allowing the plastic to conform around the patient without large gaps. It is attached to the caudal edge of the ACE with tape, and, once suction is activated, the plastic has a natural tendency to be drawn into the ACE and against all objects entering the enclosure, improving the seal. A blanket or additional weight can be added on top of the plastic over the patient to further prevent the migration of the sheet and enhance the seal.

The box was shaped to fit on a standard 66 cm (26 in) hospital stretcher while allowing both operators easy access to the patient. The ACE is 711 mm (28 in) wide at the patient’s shoulders, comfortably accommodating a male patient with shoulder width in the 99^th^ percentile with space for any tubing, lines or equipment.^14^ To better maintain a seal across beds of various sizes and to improve structural stability, two 12.7 cm (5 in) acrylic “feet” were attached along the bottom edge of the lateral walls of the box. The roof of the ACE was sloped to provide optimal visualization and space for intubation. There is adequate space to ramp the patient’s head and shoulders to align the tragus and sternum prior to intubation. This configuration provides adequate space for manipulation of airway equipment, including suction catheters, bag-valve-masks (BVM), tubes, laryngoscopes, and a bougie. Detailed schematics may be found in the supporting information.

The enclosure was designed to be cleaned between patients with sprayed 17.2% isopropyl alcohol solution, then wiped with 0.5% w/w hydrogen peroxide wipes to prevent contaminant accumulation and visual obscuration. The materials used were chosen based on their safety profile and ability to be decontaminated with commonly available agents used in hospitals.

The ability of the ACE to contain aerosols was assessed both quantitatively and qualitatively. Water/glycerol vapor smoke testing allowed for visualization of airflow around the perimeter of the ACE and through the arm ports. This smoke test was also visualized under film by a Fujifilm XT2 camera at 60 fps. The laminar movement of smoke over time through an open ACE port was measured to determine airflow velocity.

An Omega HHP886U Differential Manometer (Omega Engineering) was used to quantify the negative pressure between the ambient atmosphere and the internal ACE atmosphere. The internal pressure gauge tubing was fed into the ACE under the box along the stretcher mattress. This test was conducted under multiple conditions: (1) one operator using two ports, (2) two operators using all four ports, (3) two operators performing a simulated video laryngoscopy facilitated intubation and (4) two operators performing simulated BVM ventilation. Procedures were conducted using a CRiSis Manikin™ (GTSimulators) to simulate a patient. These trials were all conducted with 15 L min^-1^ O_2_ introduced to the ACE via standard hospital wall oxygen tubing.

Sodium Saccharin Sensitivity Test Solution (3M) for N95 respirator fit testing was used for testing the containment of aerosols in the ACE. Between five and fifty sprays of sodium saccharin were used in various test scenarios (Table 1). Assessors were exposed to a positive control of sodium saccharin and negative control of H_2_O. These assessors were blinded from the use of a negative control solution, containing only H_2_O, versus sodium saccharin solution. They were positioned 30 cm from an open port and asked if they could taste the solution under different test conditions (Table 1). These trials were all conducted with 15 L min^-1^ O_2_ being introduced to the ACE via standard hospital wall oxygen tubing. The results were analyzed by Cochran’s Q test in SPSS™ (IBM).

**Table 1:** FT-12 Sodium Saccharin Fit Test.

| Scenario | Vac.<sup>a</sup> | Assessor |  |  |  |
| --- | --- | --- | --- | --- | --- |
|  |  | 1 | 2 | 3 | 4 |
| Positive control<sup>b</sup> | N/A | Y | Y | Y | Y |
| No ports in use | On | N | N | N | N |
| No ports in use (50 sprays) | On | N | N | N | N |
| All ports in use, users stationary | On | N | N | N | N |
| All ports in use, simulated bagging | On | N | N | N | N |
| All ports in use, simulated intubation | On | N | N | N | N |
| Negative control<sup>b</sup> | Off | N | N | N | N |
| No ports in use | Off | Y | N | N | N |
| No ports in use (25 sprays) | Off | Y | Y | Y | N |

Y/N = Sodium saccharin detected by subject (yes/no).
**a**: Flow out of the enclosure with vacuum engaged was 70 L min<sup>-1</sup>.
- b:** Direct spray to subject with FT-11 Sodium Saccharin Fit Test Solution (low concentration). **c:** Negative control performed with H<sub>2</sub>O with no ports in use.

To further test aerosol containment, fluorescein solution was atomized with a MADgic Atomizer™ (Teleflex) attached to a 10 mL syringe. The atomizer was placed on the stretcher inside the ACE, 15 cm from a port with the nozzle directed vertically. Filter paper was placed 1 cm outside the port to contain any fluorescein escaping the ACE. An ultraviolet (UV) light (λ = 365 nm) was then used over the filter paper, and photographs were taken of the fluorescein’s dispersal pattern. Cleaning and reuse were also evaluated and are described in the supporting information.

## Results

Video analysis of the vapor smoke moving linearly through an open ACE port was used to calculate the approximate inflow velocity of an air leak at unused Port 1. Smoke entering the enclosure traveled approximately 30 cm perpendicular to Port 1 over 0.33 seconds. This corresponds to approximately 0.9 m/sec inflow velocity, which is above the containment requirement of Class 2 biosafety cabinets.^12^ Photographs of smoke traveling through a port, with and without suction, are shown in Figure 4.

The pressure differential Δ*P* across the enclosure over time was measured with two ports in use by an operator, first when the remaining ports were occluded with plastic, and again when a second operator entered the enclosure (Figure 2). In both cases, a pressure differential within the ACE was achieved in < 10 seconds once all ports were in use. This pressure differential drives air inflow across small gaps around the ACE to contain aerosols. A solitary leak of up to 5 cm in diameter would still provide greater than 0.5 m/s inflow velocity.

**Figure 2.**
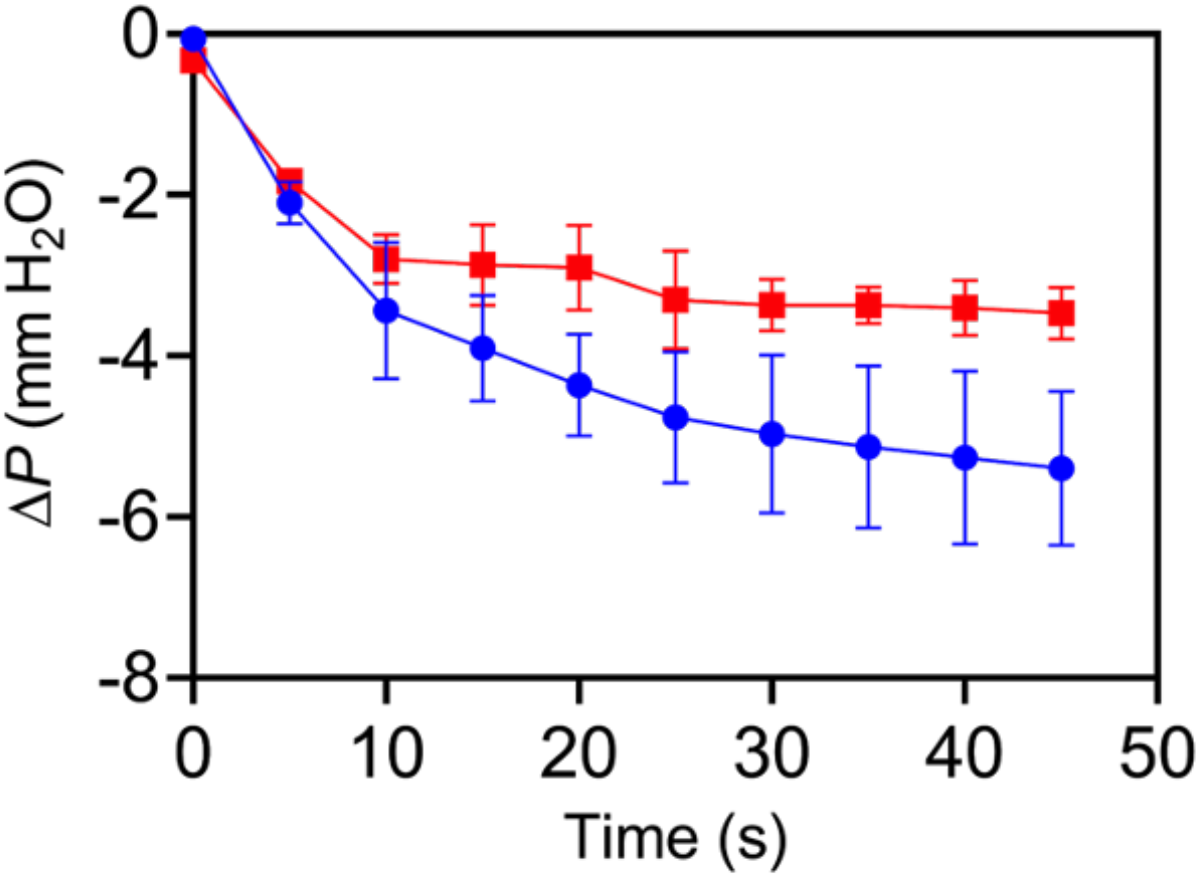
Differential pressure with front ports in use when: (red) side ports are covered with plastic sheets and (blue) a second user places his/her arms in the side ports (both users wearing recommended PPE for AGMPs). Plots are averages of three trials, with error bars indicating standard deviation.

Differential pressure was then measured during simulated BVM ventilation and endotracheal intubation scenarios. Peak pressures of -31.4 and -31.2 mm H_2_O were recorded in each scenario (Figure 3), with negative pressures maintained throughout both procedures. A less gas-permeable mattress cover was used under the box during this measurement, which resulted in a greater pressure differential.

**Figure 3.**
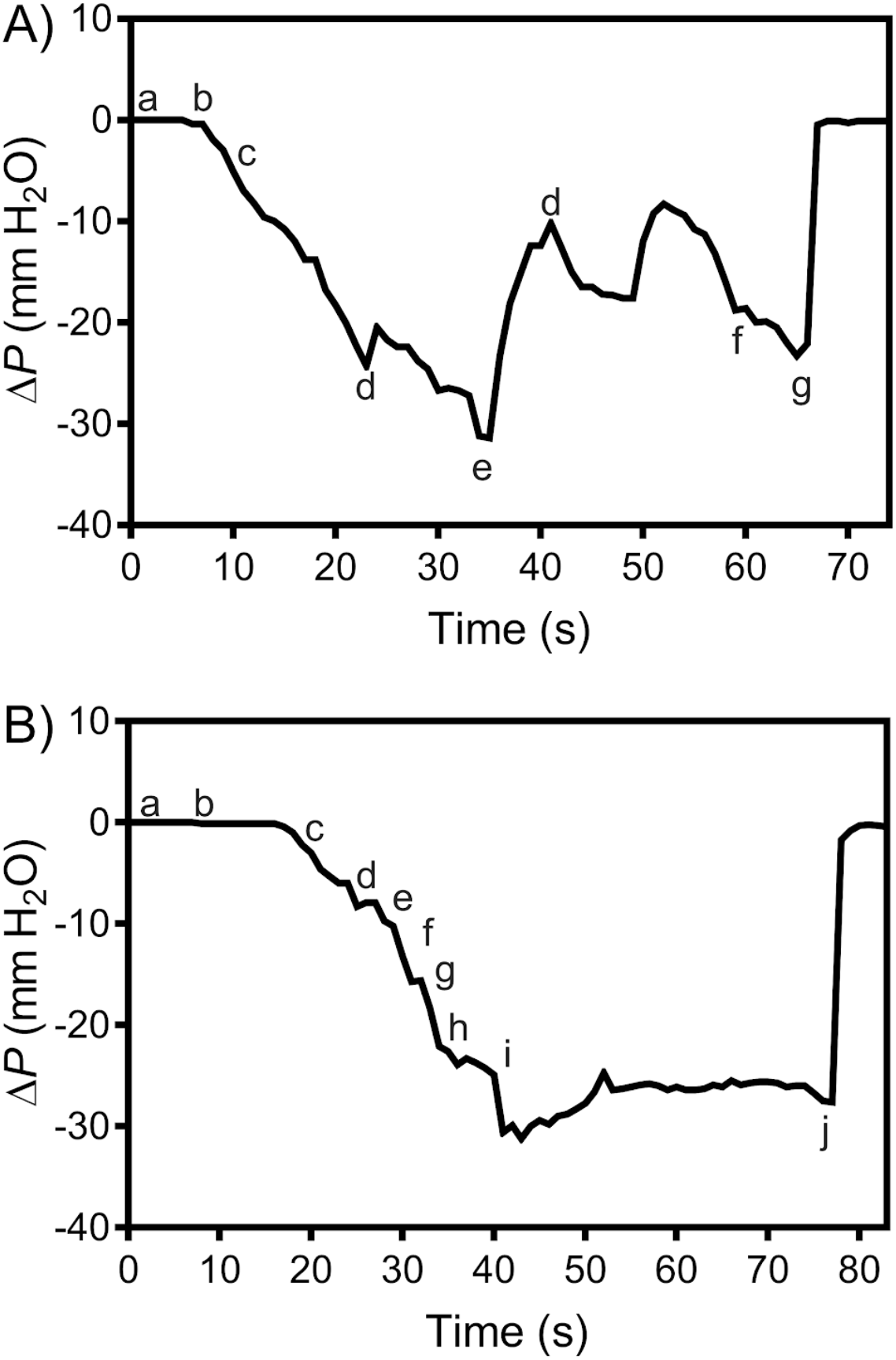
A) Differential pressure during simulated manual ventilation within the ACE with BVM at 15 L min^-1^ O_2_. **a:** Operator’s hands enter ACE ports. **b:** BVM applied to the simulated patient. **c:** Manual ventilation begins. **d:** Plastic tarp manipulated by the operator. **e:** Simulated patient repositioned by the operator. **f:** Manual ventilation stops. **g:** Operator’s hands are withdrawn. B) Differential pressure during simulated intubation using video laryngoscopy. **a:** Operator’s hands enter ACE ports. **b:** Mask applied to the simulated patient for pre-oxygenation. **c:** Mask removed. **d:** Video laryngoscopy begins. **e:** Wall suction used to simulate clearance of secretions. **f:** Endotracheal tube placed beyond vocal cords. **g:** Stylet removed from the endotracheal tube. **h:** Cuff on the endotracheal tube inflated. **i:** Endotracheal tube connected for manual ventilation. **j:** Operator’s hands withdrawn.

The silicone port gaskets allow for considerable arm maneuverability (15 cm radially, 10 cm forward/back), but can develop transient gaps during quick arm movements. Fluctuations in the pressure differential did occur due to these gaps, and also due to the expected manipulation of the plastic tarp from within the ACE during these procedures (Figure 3A).

Qualitative testing of the ACE’s ability to contain aerosols was performed with sodium saccharin and H_2_O negative control. Cochran’s Q test indicated a significant difference among the trials involving the ACE (X^2^ (7, N = 4) = 17.23, p < 0.05). None of the assessors could smell or taste the solution with the vacuum on, even with simulated intubation or bag-mask ventilation in progress, or with fifty sprays of sodium saccharin delivered (Table 1).

The testing of nebulized fluorescein demonstrated no leakage from the ACE when the vacuum is engaged and silicone gaskets are in place (Figure 4D). This stands in contrast to the ACE box without vacuum or silicone gaskets, where fluorescein visibly dispersed on the filter paper (Figure 4C).

**Figure 4.**
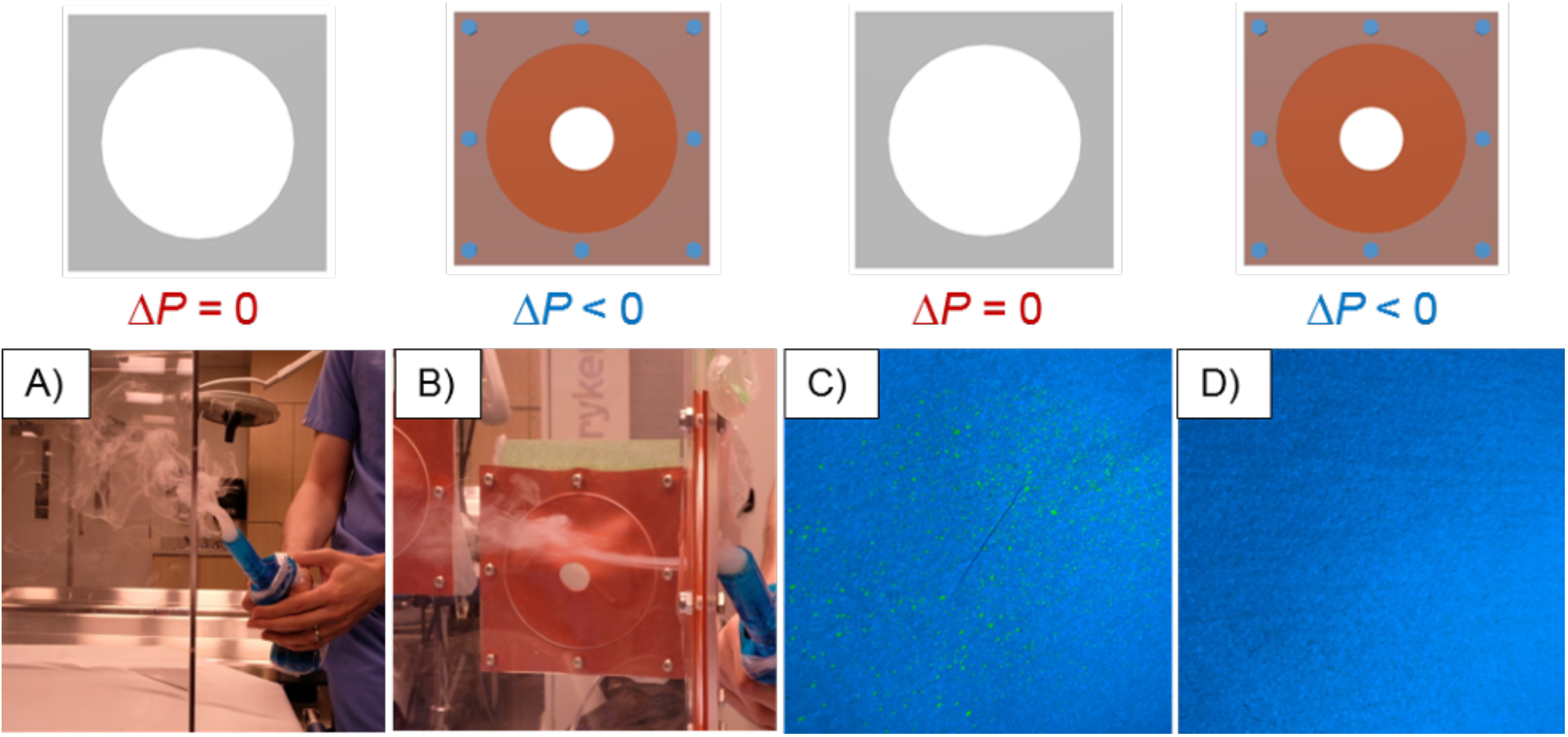
Photos depicting smoke entering enclosures with A) open ports under ambient pressure and B) ACE ports under negative pressure with schematic illustrations (top). Photos depicting fluorescein spatter on filter paper through C) open ports under ambient pressure and D) ACE ports under negative pressure.

## Discussion

The ACE maintained negative pressure in a medical simulation environment. It successfully contained smoke, saccharin and nebulized fluorescein, which suggests containment of airborne particles. Currently, there is no clear standard for shielding during AGMPs. Existing shields have been shown to decrease droplet spread, but in this testing, an open shield was not able to contain atomized fluorescein or smoke.^2^ Intubation in patients afflicted with COVID-19 presents a challenge to providers. Patients are often hypoxic prior to induction, and adequate peri-intubation oxygenation can be difficult to achieve if providers limit oxygen flows and avoid bag-valve-mask manual ventilation because of concerns about aerosolization. While some patients may tolerate transient hypoxia, others, especially those with concurrent head or spinal cord injuries, could be seriously harmed. The adoption of any type of shield is likely to increase the difficulty of first-pass intubation success. While this challenge may be reduced as the ergonomics of these shields improve, practice using the ACE in a simulation lab prior to deployment in a real scenario will be essential. The authors believe that the increased protection to providers afforded by the use of negative pressure is likely to allow for greater flexibility with oxygen delivery to patients and decrease the odds of peri-intubation hypoxia and its associated complications.

The ACE is ideally deployed in a supine or near-supine patient. It may not be suitable for patients who are agitated, claustrophobic, or whose oxygenation impairment precludes lying flat. A delayed sequence oxygenation and ventilation strategy may allow the ACE to be deployed once the patient is sedated. In the case of an unsuccessful intubation or an urgent requirement for anterior neck access, the ACE is easy to remove. The authors recommend its removal be practiced in a simulated environment, and do not currently recommend its deployment in a scenario where first-pass success may be unlikely.

## Limitations

The containment enclosure reported here has several limitations. While acrylic is both readily available and inexpensive, it is prone to cracking on impact and may shatter when dropped from a height or with a severely agitated patient. An earlier prototype fabricated from polycarbonate was found to be more durable in comparison. Setup for this protocol also requires additional time and training relative to procedures with no enclosure. The differential pressure achievable using the ACE is dependent on both the available sources of suction and the permeability of the mattress used. The ACE should only be used in facilities where the provided suction system can safely dispose of any infectious matter.

## Conclusion

The ACE was found to be effective in limiting aerosol dispersion. It is cost-effective and can be assembled without specialized tools or training. While the ACE will not obviate the need for healthcare providers to wear airborne precautions during intubation, it may further decrease viral transmission risk during AGMPs. With disposable PPE supplies currently constrained in many jurisdictions, a negative pressure enclosure such as the ACE may aid in improving protection for healthcare providers.

## Data Availability

All data collected is presented in the manuscript or supplementary index.

## Acknowledgments

The authors would like to thank David Sommer, Steven Rogak, Michael Rooney, Fraser Parlane, Benjamin MacLeod, and Karry Ocean for valuable discussions, and Kylie Perrins, Manager of the Vancouver General Hospital Simulation Centre, for providing a wet lab to conduct our tests. The authors also thank Barry Creighton and Clare Creighton for enabling testing of an early version of the enclosure.

## References

1. Tran K, Cimon K, Severn M, Pessoa-Silva CL, Conly J. Aerosol generating procedures and risk of transmission of acute respiratory infections to healthcare workers: a systematic review. PLOS ONE. 2012;7(4):e35797. doi:10.1371/journal.pone.0035797

2. Canelli R, Connor CW, Gonzalez M, Nozari A, Ortega R. Barrier enclosure during endotracheal intubation. N Engl J Med. 2020;0(0):null. doi:10.1056/NEJMc2007589

3. Lai, Hsien Yung. Aerosol boxdesign. https://sites.google.com/view/aerosolbox/design. Accessed April 5, 2020.

4. Johnston LB, Conly JM. Severe acute respiratory syndrome: what have we learned two years later? Can J Infect Dis Med Microbiol. 2004;15(6):309–312.

5. Wang D, Hu B, Hu C, et al. Clinical characteristics of 138 hospitalized patients with 2019 novel coronavirus–infected pneumonia in Wuhan, China. JAMA. 2020;323(11):1061–1069. doi:10.1001/jama.2020.1585

6. Wu Z, McGoogan JM. Characteristics of and important lessons from the coronavirus disease 2019 (covid-19) outbreak in China: summary of a report of 72 314 cases from the chinese center for disease control and prevention. JAMA. February 2020. doi:10.1001/jama.2020.2648

7. Integrated surveillance of COVID-19 in Italy. https://www.epicentro.iss.it/en/coronavirus/bollettino/Infografica_5aprile%20ENG.pdf.

8. Apr 02 LP· CN· P, April 2 2020 4:00 AM ET | Last Updated: Health-care workers make up 1 in 10 known cases of COVID-19 in Ontario | CBC News. CBC. https://www.cbc.ca/news/canada/toronto/health-care-workers-make-up-1-in-10-known-cases-of-covid-19-in-ontario-1.5518456. Published April 2, 2020. Accessed April 5, 2020.

9. Ji Y, Ma Z, Peppelenbosch MP, Pan Q. Potential association between COVID-19 mortality and health-care resource availability. Lancet Glob Health. 2020;8(4):e480. doi:10.1016/S2214-109X(20)30068-1

10. World Health Organization, Pandemic and Epidemic Diseases, World Health Organization. Infection Prevention and Control of Epidemic-and Pandemic-Prone Acute Respiratory Infections in Health Care: WHO Guidelines.; 2014. http://apps.who.int/iris/bitstream/10665/112656/1/9789241507134_eng.pdf?ua=1. Accessed April 5, 2020.

11. van Doremalen N, Bushmaker T, Morris DH, et al. Aerosol and surface stability of SARS-CoV-2 as compared with SARS-CoV-1. N Engl J Med. 2020;0(0):null. doi:10.1056/NEJMc2004973

12. National Sanitation Foundation. NSF/ANSI 49-2019: Biosafety Cabinets -Design, Construction, Performance, And Field Certification. March 2020. https://www.techstreet.com/nsf/subgroups/209. Accessed April 5, 2020.

13. Feng X, Zhang Y, Xu Z, Song D, Cao G, Liang L. Aerosol containment by airflow in biosafety laboratories. J Biosaf Biosecurity. 2019;1(1):63–67. doi:10.1016/j.jobb.2018.12.009

14. Robert Lewis, ed. Human Integration Design Handbook. Revision 1. Washington, DC 20546-0001: National Aeronautics and Space Administration; 2014. https://www.nasa.gov/feature/human-integration-design.

